# Mortality Trends in the United States due to Concurrent Heart Failure, Atrial Fibrillation/Flutter, and Sepsis

**DOI:** 10.64898/2026.02.06.26345787

**Authors:** Gaithrri Shanmuganathan, Nicole Jeongha Lee, Devendra K. Agrawal

## Abstract

**Background:** Heart failure (HF), atrial fibrillation (AF)/atrial flutter (AFL) and sepsis commonly co-occur in hospitalized patients. This study determines temporal mortality trends associated with concurrent HF, AF/AFL, and sepsis among adults across demographic and geographic groups in the United States.

**Methods:** The CDC Wonder database was utilized to extract age-adjusted mortality rates (AAMR) per 100,000 for deaths listing HF, AF/AFL, and sepsis. Trends were analyzed by age, race/ethnicity, region, and sex. Joinpoint regression calculated the annual percent change (APC) and average annual percent change (AAPC) for AAMR with 95% CI.

**Results:** From 1999 to 2023, there were 1,749,565 deaths involving HF, AF/AFL, and sepsis. AAMR doubled (1999: 11.79 vs 2020: 23.87 per 100,000), with a critical 2012 inflection point accelerating mortality from 1.29% to 6.42% annually. White individuals had steepest post-2012 acceleration (6.67%), surpassing Black individuals by 2020 (24.88 vs 20.80 per 100,000). Males had higher AAMRs than females (28.69 vs 20.19 per 100,000 in 2020). Middle-aged adults (45-64 years) showed highest acceleration (9.98-10.30%), nearly double those aged ≥85 years (5.82%). The Midwest and South had steepest increases (7.07% and 7.11%). During 2018–2023, mortality continued increasing at 6.11% annually without stabilization.

**Conclusions:** Mortality involving HF, AF/AFL and sepsis doubled from 1999–2023 with sustained acceleration and no post-pandemic stabilization. Targeted interventions should focus on males, middle-aged adults, and high-risk regions with enhanced post-discharge care.

**Project Overview:**

- This is retrospective cohort study using CDC WONDER data (1999-2023).
- We analyzed 1,749,565 deaths involving all three conditions, heart failure, atrial fibrillation/flutter, and sepsis.
- To our knowledge, this is the first population-level study examining mortality trends due to the coexistence of heart failure, atrial fibrillation/flutter, and sepsis.

**Key Findings:**

- verall age-adjusted mortality rates doubled from 11.79 to 23.87 per 100,000 (1999-2020).
- was a critical inflection point in 2012 with marked acceleration thereafter.
- findings revealed significant demographic disparities regarding age, sex, race/ethnicity, and region.
- acceleration through 2023 was found without post-pandemic. stabilization.

## 1. Introduction

Heart failure (HF) affects more than 7.4 million adults in the United States and remains a leading cause of hospitalization and mortality [1]. Atrial fibrillation (AF) is the most common sustained cardiac arrhythmia, with prevalence projected to reach 12.1 million by 2030 [2]. Atrial flutter (AFL) frequently coexists with AF [3]. Sepsis contributes to 1.7 million hospitalizations and 350,000 deaths annually [4]. When all three conditions coexist, medical management becomes particularly complex as each condition can exacerbate the others. Previous studies have investigated mortality trends in dyads involving two of these conditions. However, no studies have examined mortality trends when all three conditions coexist, leaving a critical knowledge gap. We hypothesize that mortality in patients with concurrent HF, AF/AFL, and sepsis has increased over time with temporal acceleration in the early 2010s following healthcare policy changes, and heterogeneous trends across different demographics and geographic regions. In this study, we describe the long-term (1999–2020) and recent (2018–2023) temporal trends in mortality in the United States across different demographics and geographic groups.

## 2. Methods

### 2.1. Study Design and Database

The CDC Wide-Ranging Online Data for Epidemiologic Research (WONDER) Multiple Cause of Death (MCOD) database was analyzed from January 1, 1999 to December 31, 2023 in adults ≥ 25 years to conduct this retrospective, population-based analysis of mortality trends [5]. Deaths for HF, AF/AFL, and sepsis were identified using the International Classification of Diseases, Tenth Revision (ICD-10) codes. We applied a multiple cause-of-death approach in which conditions are listed anywhere on the death certificate. HF cases were defined using codes I11.0 (hypertensive heart disease with heart failure), I13.0 (hypertensive heart and renal disease with heart failure), I13.2 (hypertensive heart and renal disease with both heart failure and renal failure), and I50.x (all heart failure subtypes). AF/AFL cases were identified using code I48.x (all AF and AFL subtypes). Sepsis was identified using codes A40.x (streptococcal sepsis) and A41.x (other sepsis). Institutional Review Board (IRB) approval was not required as this study utilized de-identified, publicly available data. The author team includes representation across genders, institutional backgrounds and career stages.

### 2.2. Statistical Analysis

The primary outcome was the age-adjusted mortality rate (AAMR) per 100,000 population, calculated using the direct method and standardized to the 2000 US standard population [6]. Crude mortality rates were calculated for age-stratified descriptive analyses. Joinpoint regression analysis (Joinpoint Regression Program, Version 5.0.2, Statistical Research and Applications Branch, National Cancer Institute) was used to assess temporal trends in mortality rates [7]. This program identifies statistically significant changes in the slope of log-transformed rates over time. To determine the optimal number and location of joinpoints, models were fitted using the modified Bayesian Information Criterion (mBIC). Separate analyses were performed stratified by age group (25–34, 35–44, 45–54, 55–64, 65–74, 75–84, and ≥85 years), sex, race/ethnicity (White, Black, Asian/Pacific Islander, American Indian/Alaska Native, and Hispanic), and United States census region (Northeast, Midwest, South, and West). Results are reported as annual percent change (APC) for individual segments and average annual percent change (AAPC) for the full interval, along with their 95% confidence intervals. Statistical significance was defined as P<0.05.

## 3. Results

### 3.1 Overall Temporal Trends

From 1999 to 2020, there were 1,152,066 deaths involving HF, AF/AFL, and sepsis in the United States. AAMR increased significantly from 11.79 per 100,000 in 1999 to 23.87 per 100,000 in 2020. Joinpoint regression identified one statistically significant inflection point in 2012, resulting in two distinct temporal segments. Between 1999 and 2012, mortality increased significantly at an APC of 1.29% (95% CI, 0.85%–1.65%; p<0.001). Following 2012, the rate of increase accelerated markedly, with mortality rising at an APC of 6.42% per year from 2012 to 2020 (95% CI, 5.64%–7.46%; p<0.001). Overall, the AAPC for the full 1999–2020 period was 3.21% (95% CI, 2.99%–3.43%), indicating a sustained and statistically significant increase in mortality across the study period (**Table 1**, **Figure 1**).

**Figure 1:**
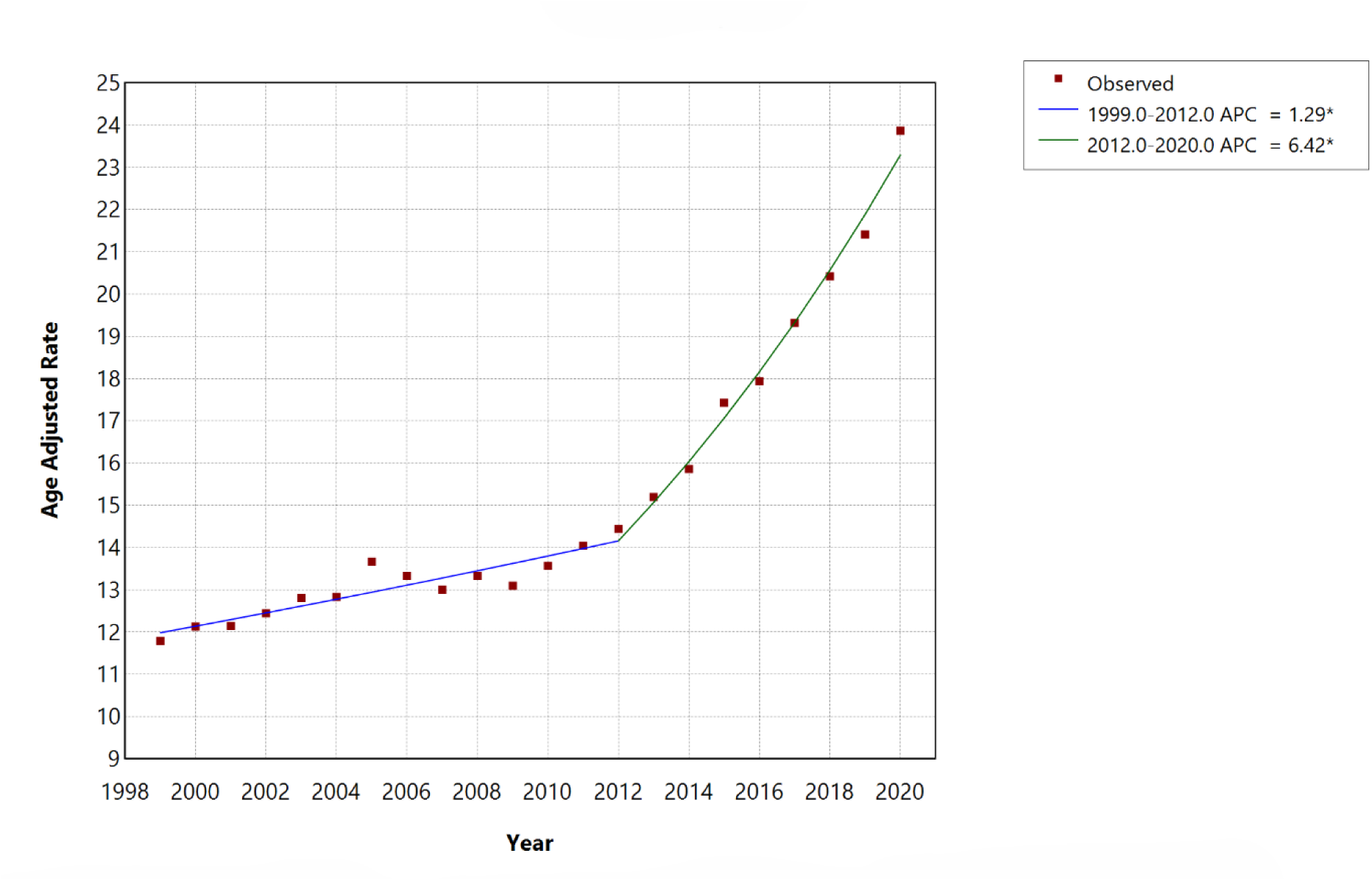
Overall HF, AF/AFL and sepsis-related Age-Adjusted Mortality Rates per 100,000 among individuals aged ≥25 years in the United States, 1999 to 2020. ∗ Indicates that the annual percentage change (APC) is significantly different from zero at α=0.05. APC=annual percent change.

**Table 1:**
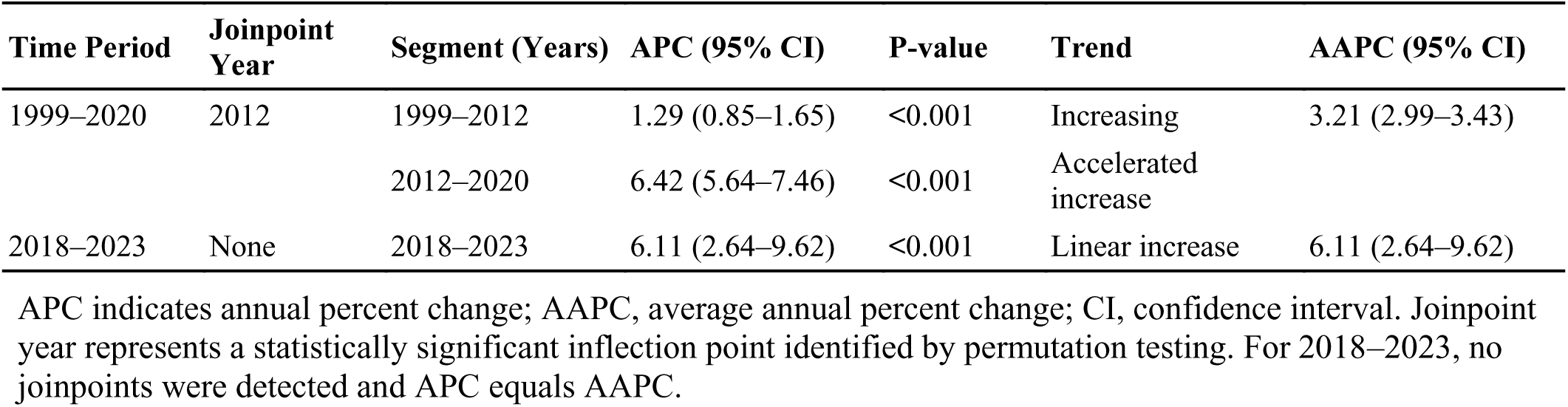
Overall Joinpoint Trends in Age-Adjusted Mortality Rates Associated with Heart Failure, Atrial Fibrillation/Flutter, and Sepsis in the United States.

During the 2018–2023 period, joinpoint analysis identified no statistically significant joinpoints, and mortality trends were best described by a single linear segment. AAMR increased from 20.42 to 26.69 per 100,000, continuing to increase significantly at an APC of 6.11% per year (95% CI, 2.64%–9.62%; p<0.001), with the APC equivalent to the AAPC over this interval, indicating sustained acceleration without the stabilization observed for some cardiovascular conditions.

### 3.2 Race/Ethnicity-Stratified Trends

Stratified joinpoint analyses for race/ethnicity demonstrated heterogeneous trends in AAMR (**Table 2**). From 1999 to 2020, White, Black, and Hispanic populations exhibited statistically significant inflection points in 2012. Among White individuals, AAMR increased from 11.83 per 100,000 in 1999 to 24.88 per 100,000 in 2020, with mortality increasing modestly from 1999–2012 (APC 1.50%, 95% CI 1.09–1.83, p<0.001) followed by a marked acceleration from 2012–2020 (APC 6.67%, 95% CI 5.94–7.63, p<0.001), yielding an overall AAPC of 3.44% (95% CI 3.23–3.63). Black individuals demonstrated AAMR of 12.17 per 100,000 in 1999 rising to 20.80 per 100,000 in 2020, with stable mortality prior to 2012 (APC −0.38%, 95% CI −1.27 to 0.29, p=0.230) and a significant post-2012 acceleration (APC 6.08%, 95% CI 4.68–8.25, p<0.001), corresponding to an AAPC of 2.03% (95% CI 1.64–2.44). Among Hispanic individuals, AAMR increased from 7.73 per 100,000 in 1999 to 14.46 per 100,000 in 2020, with mortality remaining stable before 2012 (APC 0.62%, 95% CI −0.14 to 1.21, p=0.085) and increasing sharply thereafter (APC 6.11%, 95% CI 4.91–7.89, p<0.001), with an overall AAPC of 2.68% (95% CI 2.33–3.05) (**Table 2**, **Figure 2**).

**Figure 2:**
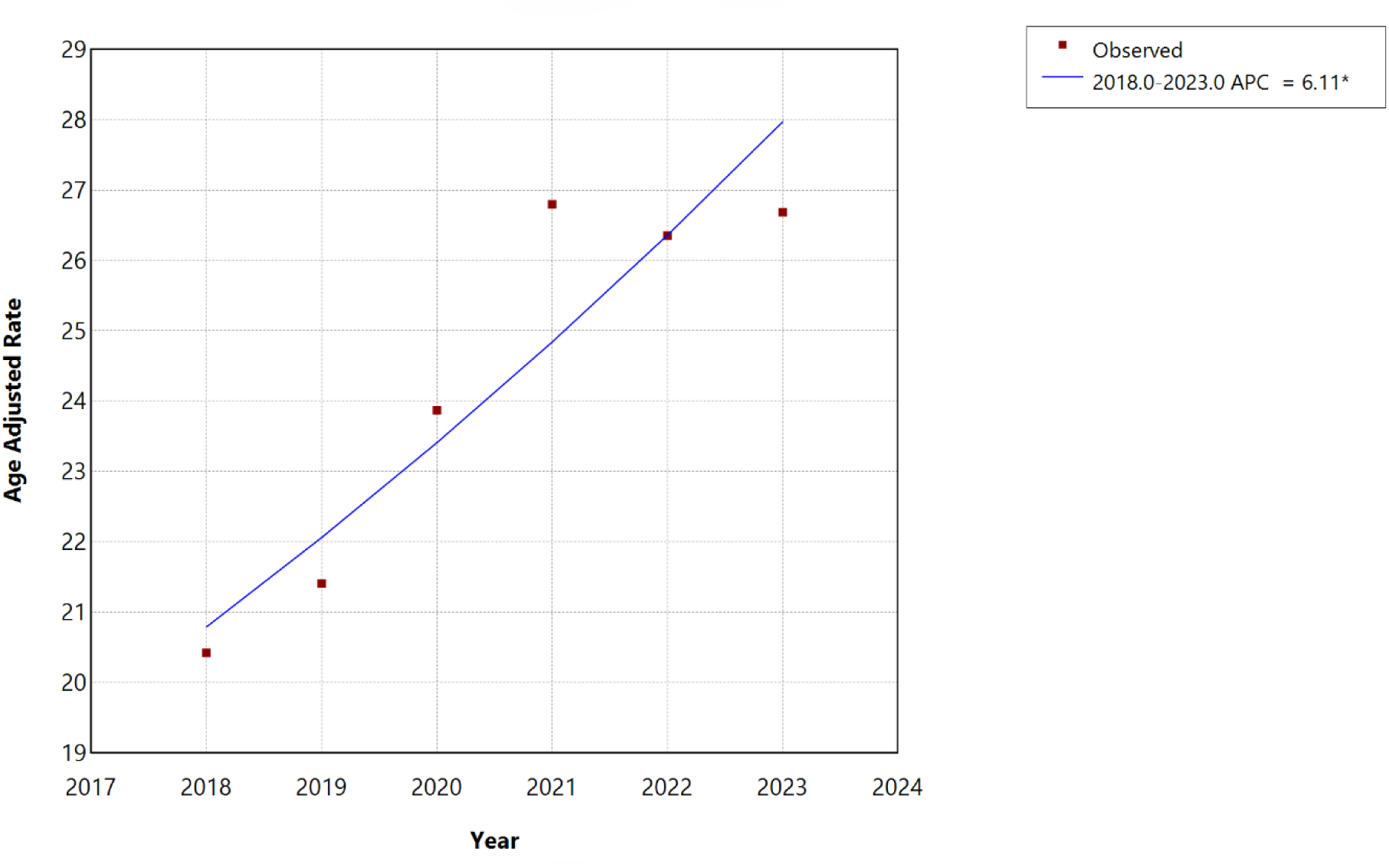
Overall HF, AF/AFL and sepsis-related Age-Adjusted Mortality Rates per 100,000 among individuals aged ≥25 years in the United States, 2018 to 2023. ∗ Indicates that the annual percentage change (APC) is significantly different from zero at α=0.05. APC=annual percent change.

**Figure 3:**
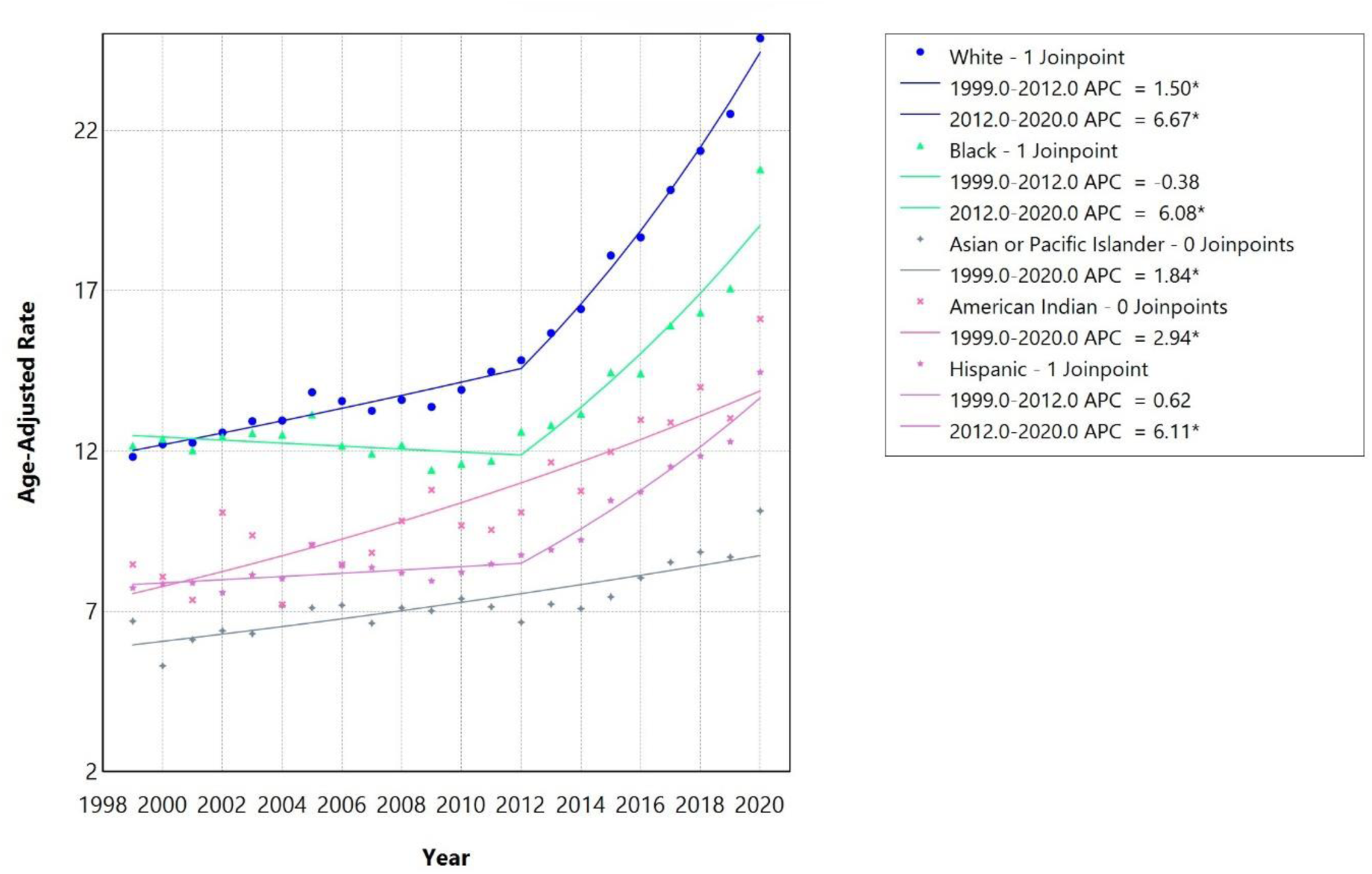
Race-stratified HF, AF/AFL and sepsis-related Age-Adjusted Mortality Rates per 100,000 among individuals aged ≥25 years in the United States, 1999 to 2020. ∗ Indicates that the annual percentage change (APC) is significantly different from zero at α=0.05. APC=annual percent change.

**Figure 4:**
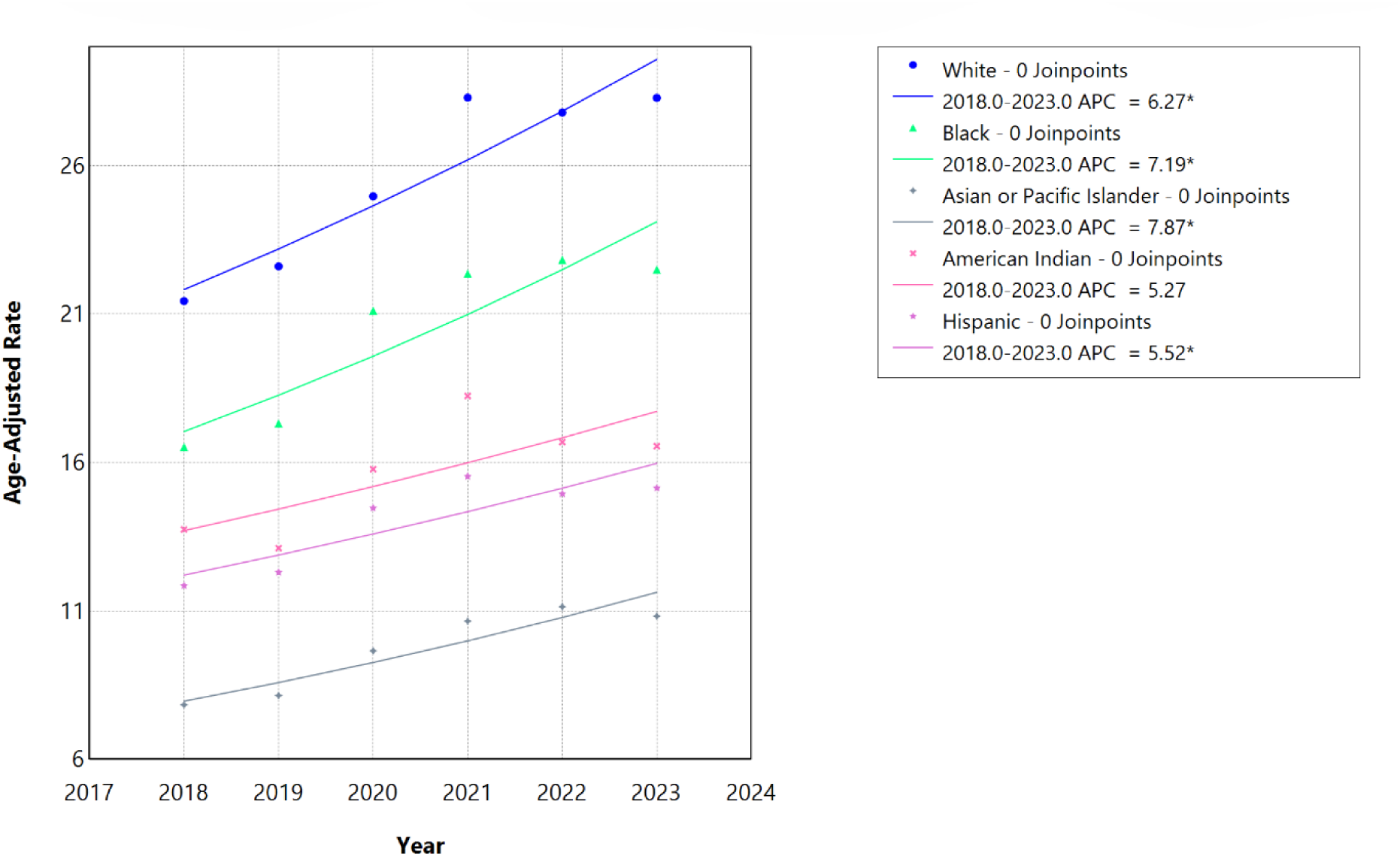
Race-stratified HF, AF/AFL and sepsis-related Age-Adjusted Mortality Rates per 100,000 among individuals aged ≥25 years in the United States, 2018 to 2023. ∗ Indicates that the annual percentage change (APC) is significantly different from zero at α=0.05. APC=annual percent change.

**Figure 5:**
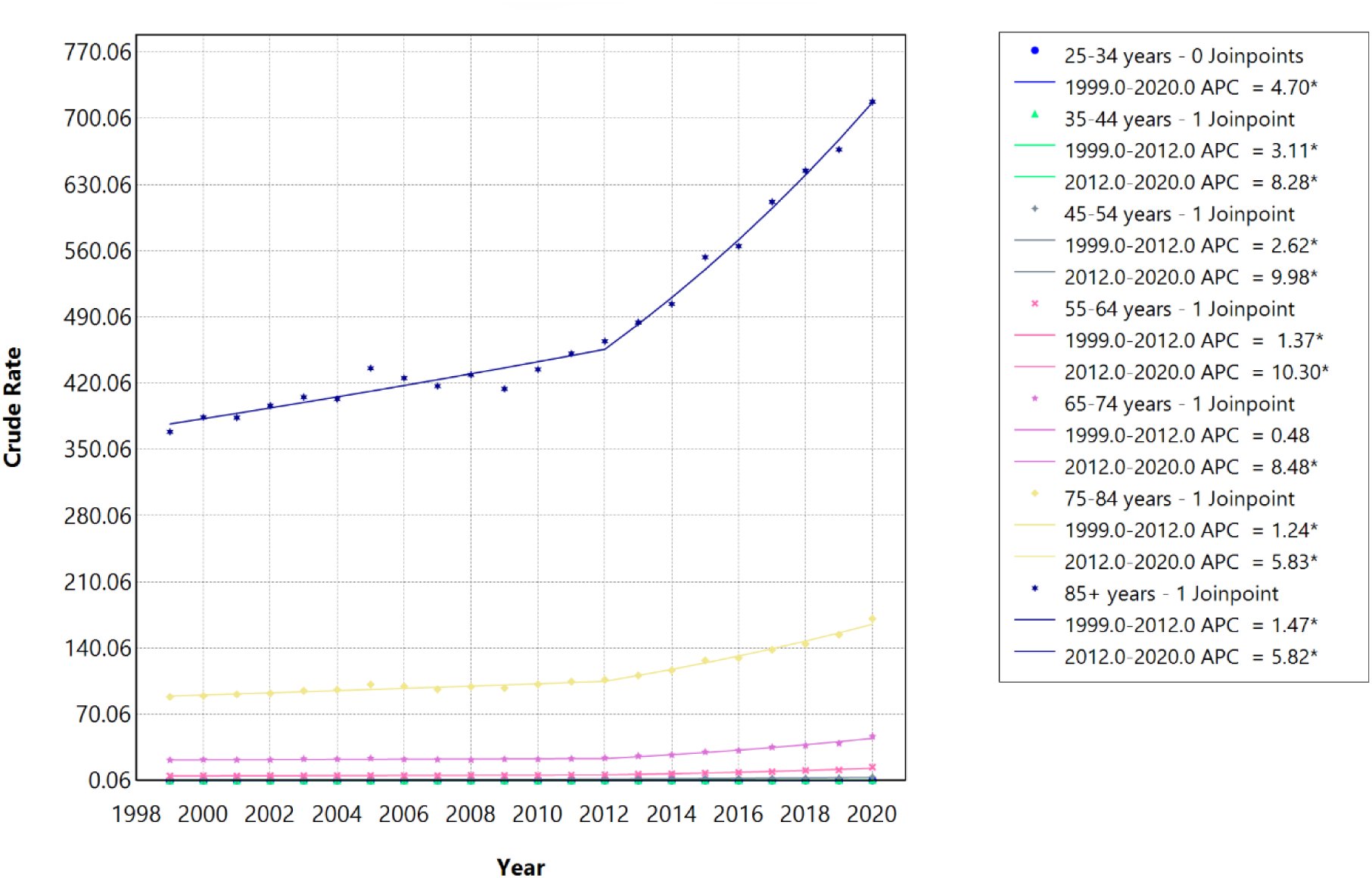
Age-stratified HF, AF/AFL and sepsis-related Crude Rates among individuals aged ≥25 years in the United States, 1999 to 2020. ∗ Indicates that the annual percentage change (APC) is significantly different from zero at α=0.05. APC=annual percent change.

**Figure 6:**
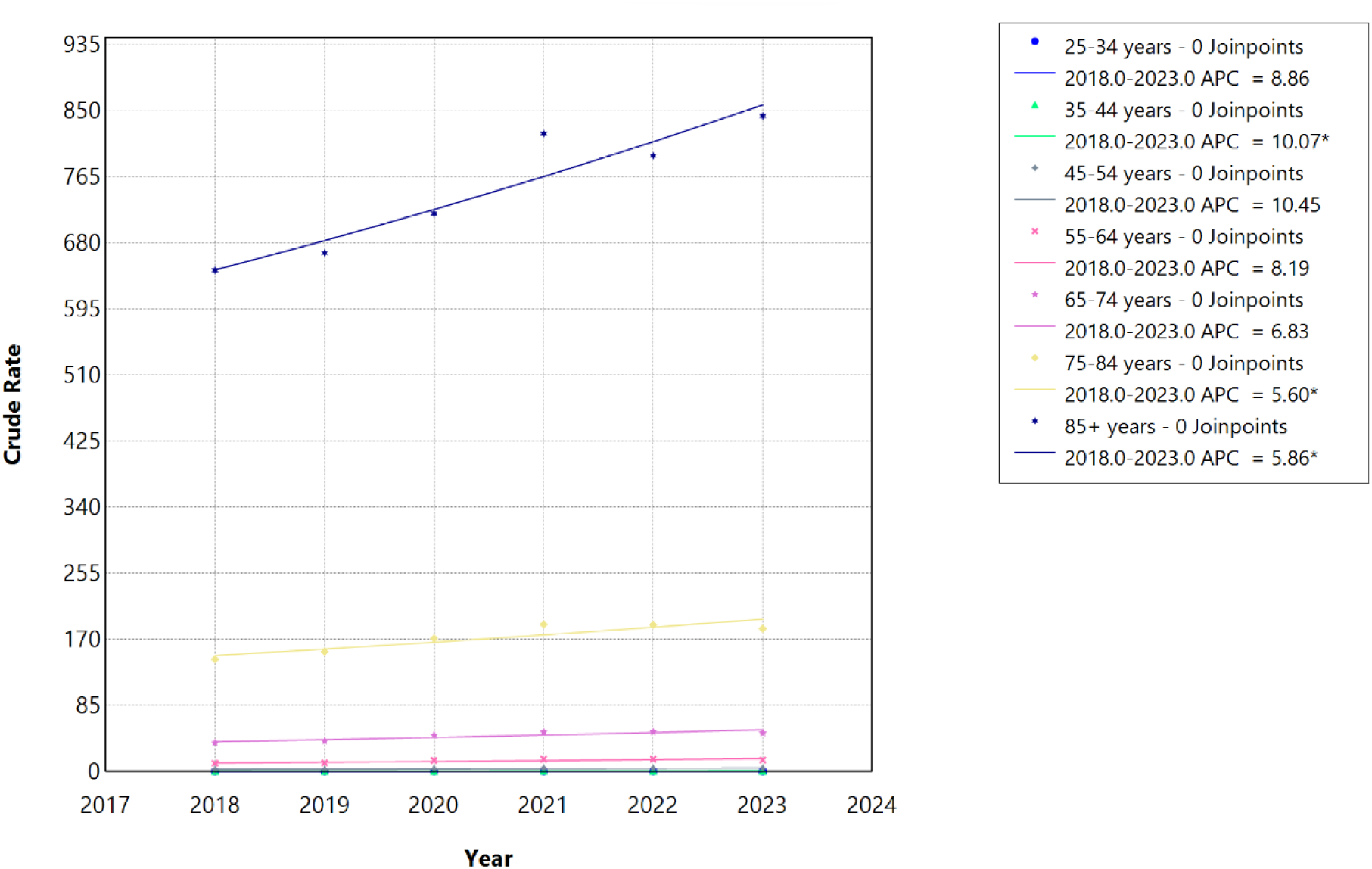
Age-stratified HF, AF/AFL and sepsis-related Crude Rates among individuals aged ≥25 years in the United States, 2018 to 2023. ∗ Indicates that the annual percentage change (APC) is significantly different from zero at α=0.05. APC=annual percent change.

**Figure 7:**
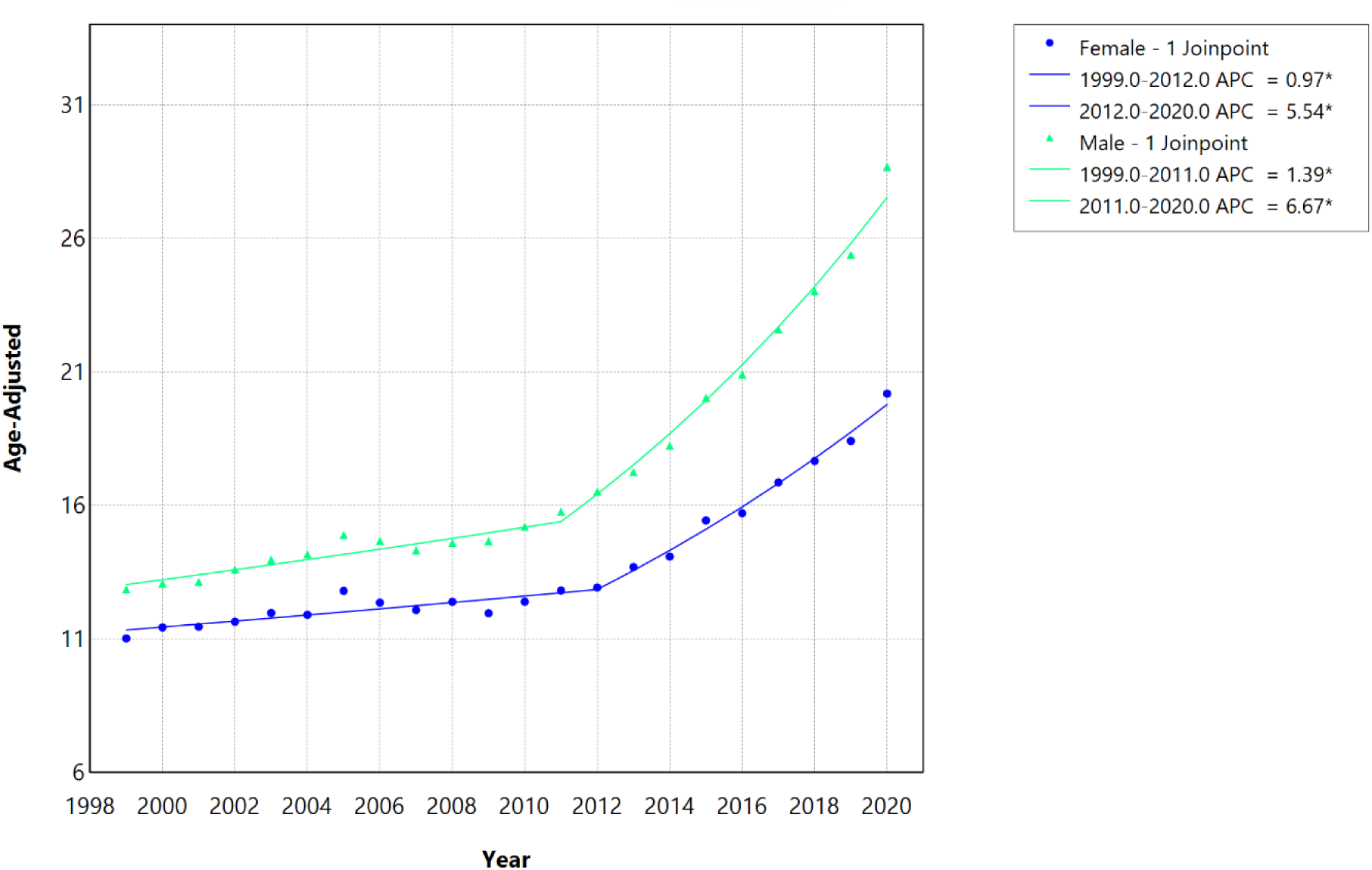
Sex-stratified HF, AF/AFL and sepsis-related Age-Adjusted Mortality Rates per 100,000 among individuals aged ≥25 years in the United States, 1999 to 2020. ∗ Indicates that the annual percentage change (APC) is significantly different from zero at α=0.05. APC=annual percent change.

**Figure 8:**
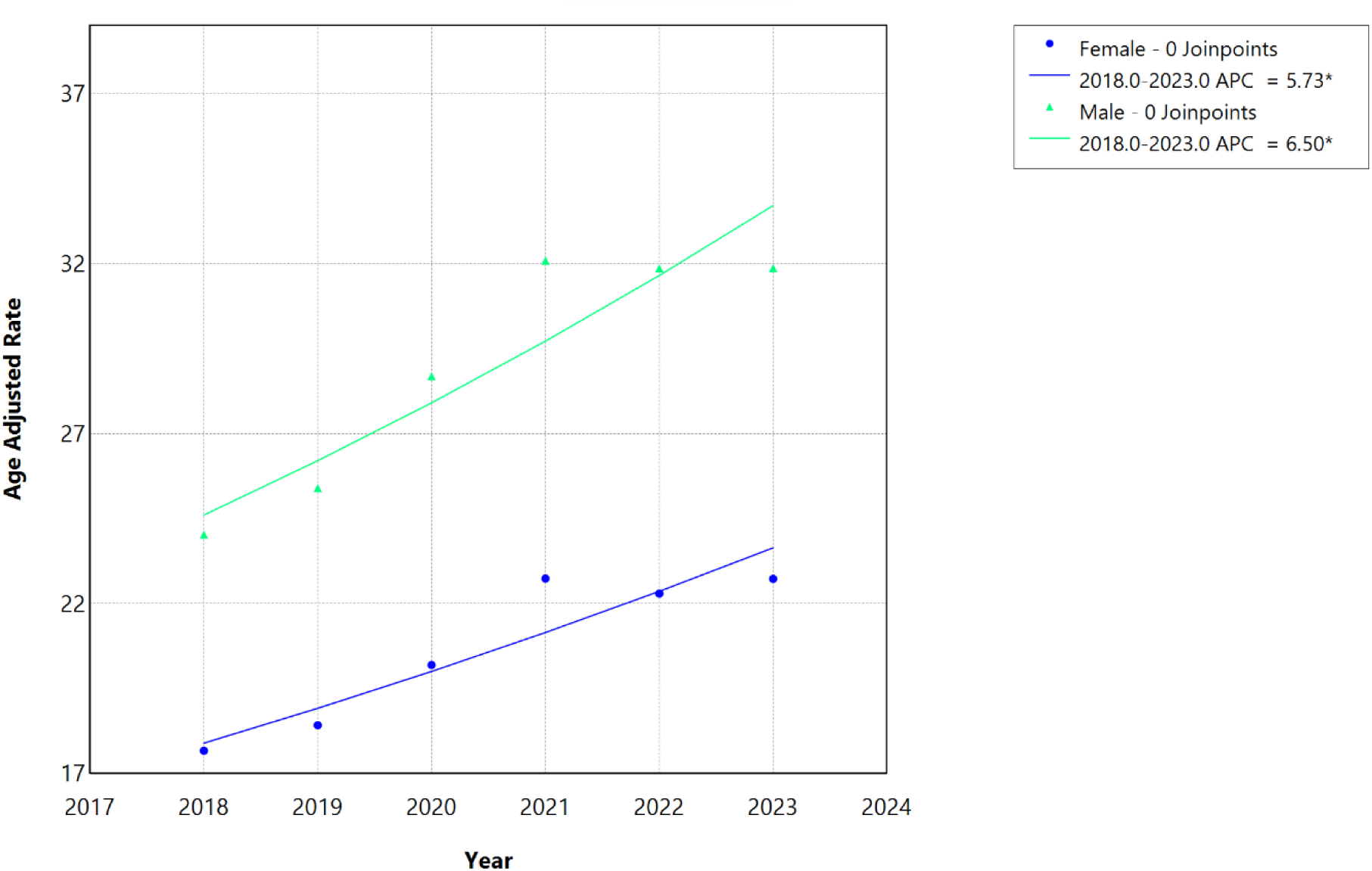
Sex-stratified HF, AF/AFL and sepsis-related Age-Adjusted Mortality Rates per 100,000 among individuals aged ≥25 years in the United States, 2018 to 2023. ∗ Indicates that the annual percentage change (APC) is significantly different from zero at α=0.05. APC=annual percent change.

**Figure 9:**
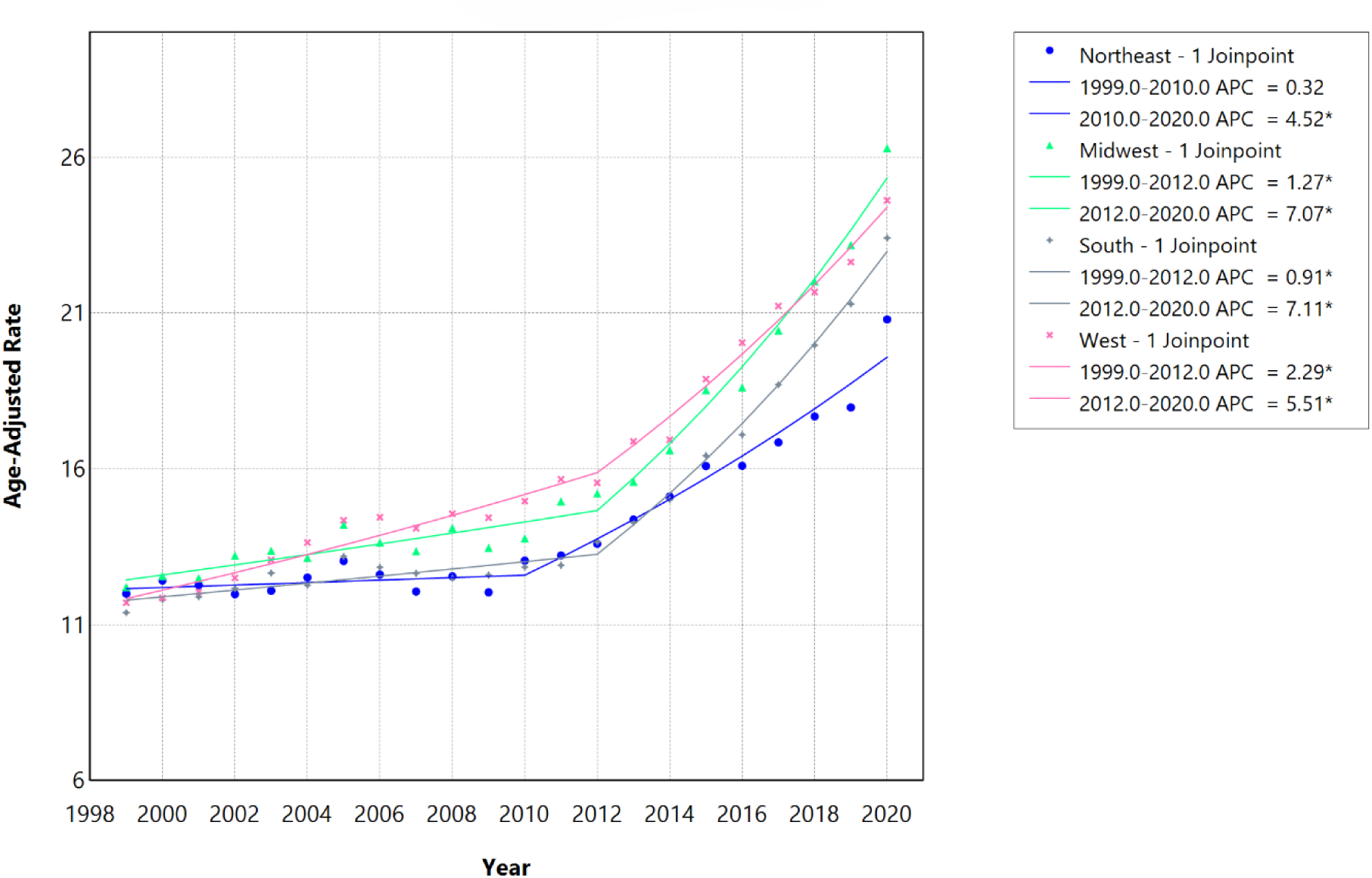
Census Region-stratified HF, AF/AFL and sepsis-related Age-Adjusted Mortality Rates per 100,000 among individuals aged ≥25 years in the United States, 1999 to 2020. ∗ Indicates that the annual percentage change (APC) is significantly different from zero at α=0.05. APC=annual percent change.

**Figure 10:**
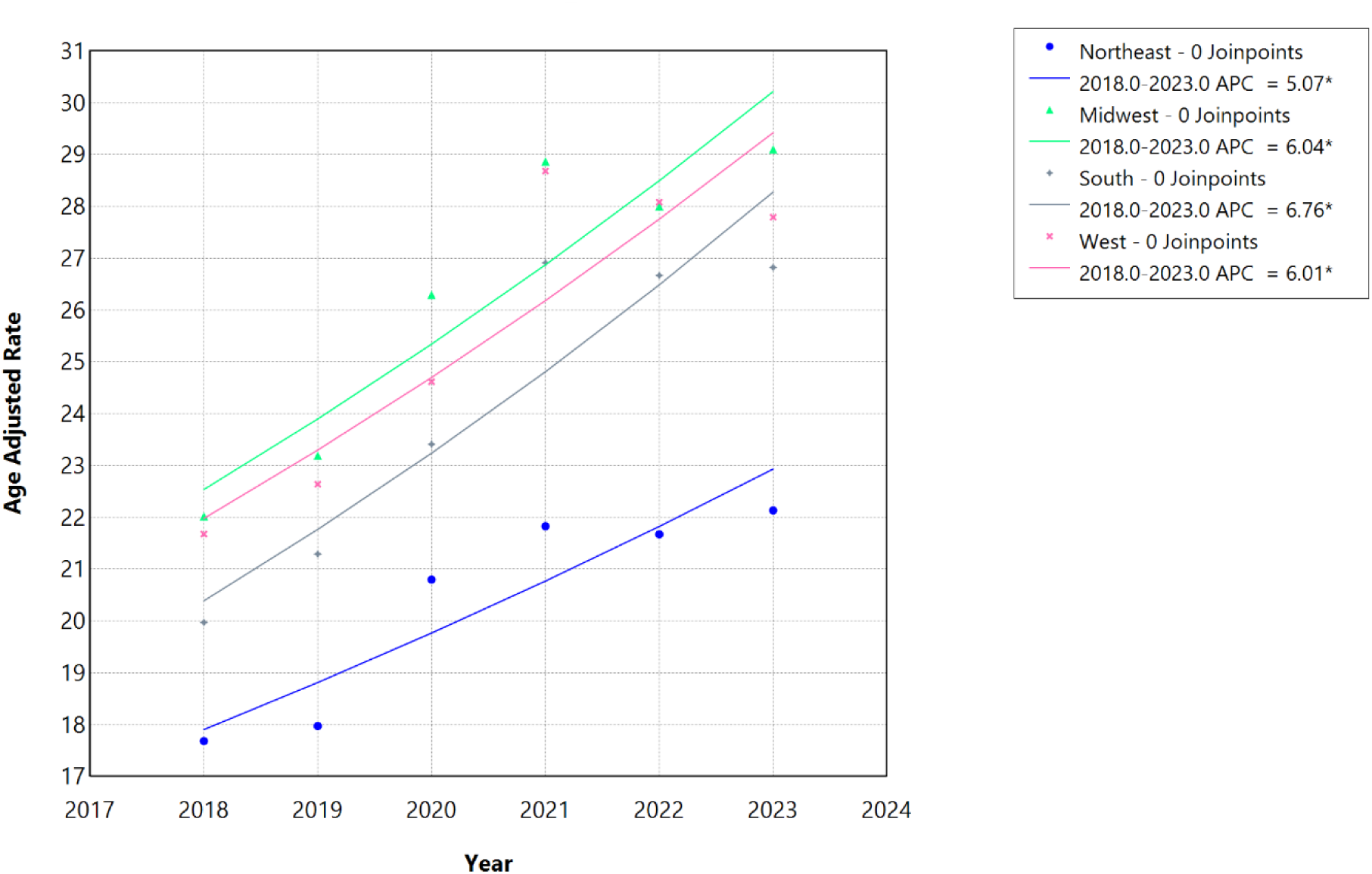
Census Region-stratified HF, AF/AFL and sepsis-related Age-Adjusted Mortality Rates per 100,000 among individuals aged ≥25 years in the United States, 2018 to 2023. ∗ Indicates that the annual percentage change (APC) is significantly different from zero at α=0.05. APC=annual percent change.

**Table 2:**
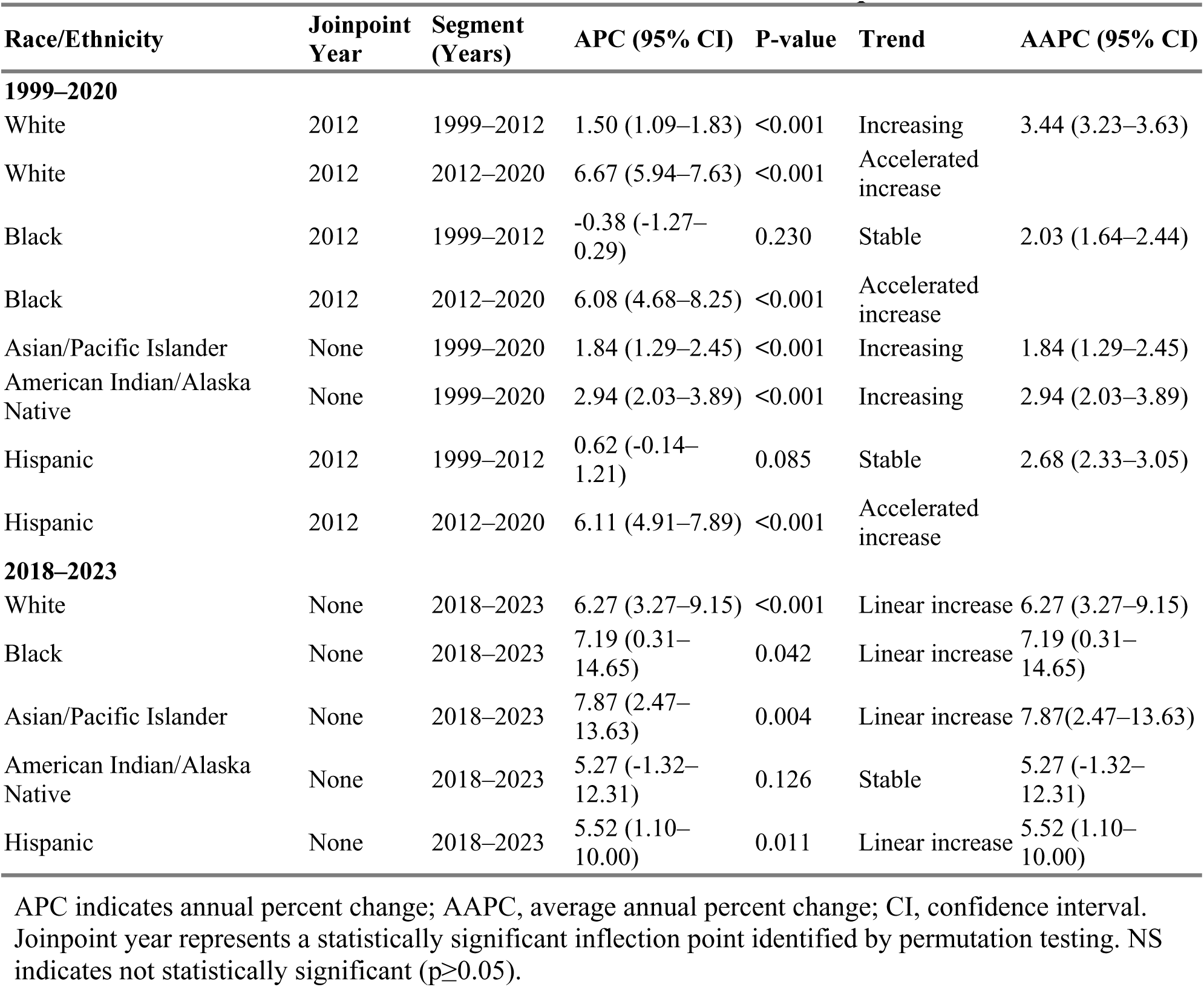
Race/Ethnicity-Specific Joinpoint Trends in Age-Adjusted Mortality Rates Associated with Heart Failure, Atrial Fibrillation/Flutter, and Sepsis in the United States.

**Table 3.**
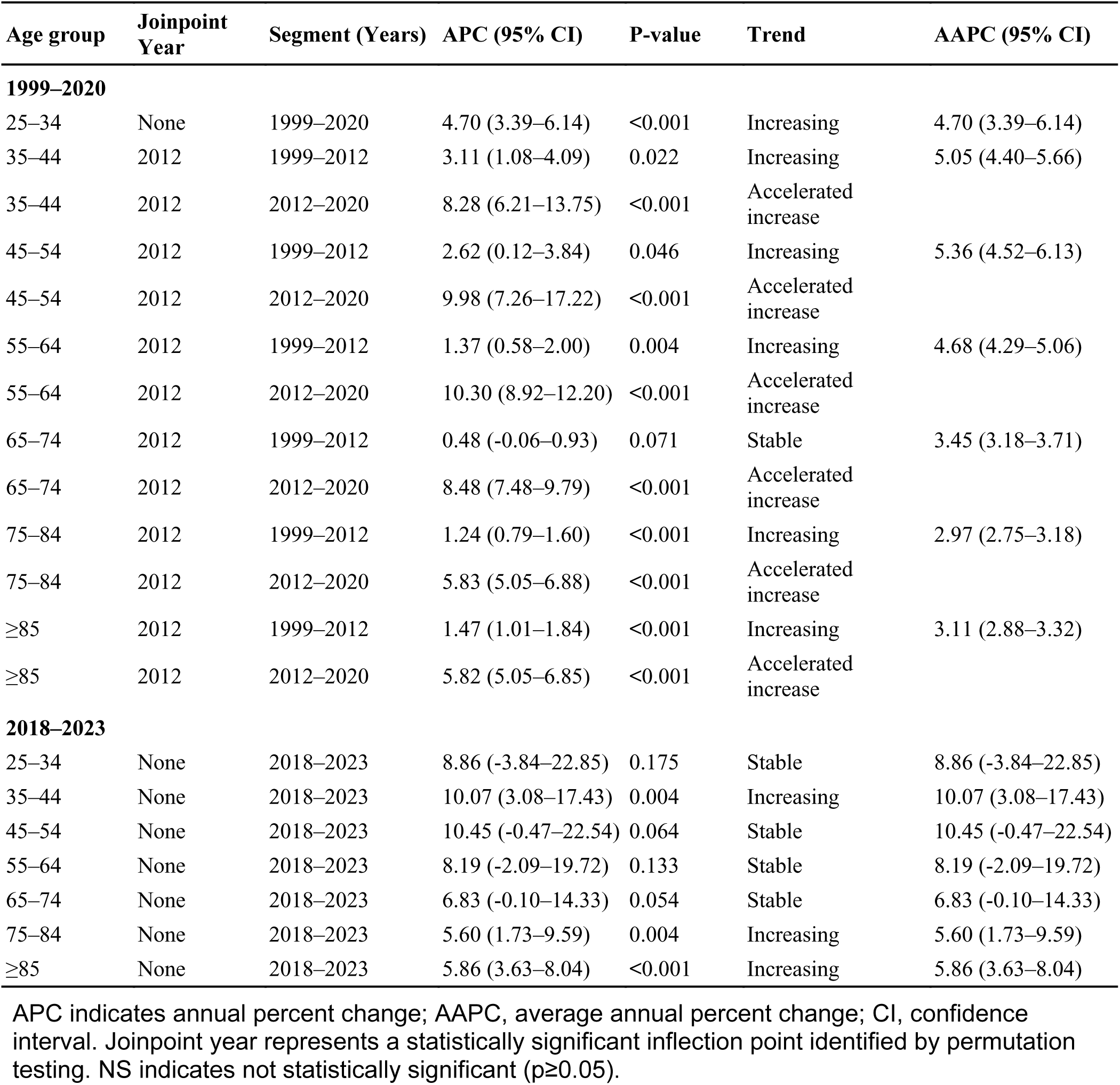
Age-Specific Joinpoint Trends in Crude Mortality Rates Associated With Heart Failure, Atrial Fibrillation/Flutter, and Sepsis in the United States.

In contrast, statistically insignificant joinpoints were detected among American Indian/Alaska Native or Asian/Pacific Islander populations. Asian/Pacific Islander individuals showed AAMR of 6.70 per 100,000 in 1999 increasing to 10.14 per 100,000 in 2020, with mortality increasing steadily across the full study period (AAPC 1.84%, 95% CI 1.29–2.45, p<0.001). American Indian/Alaska Native individuals demonstrated AAMR of 8.47 per 100,000 in 1999 rising to 16.13 per 100,000 in 2020, with steady increases throughout the period (AAPC 2.94%, 95% CI 2.03–3.89, p<0.001).

During the 2018–2023 period, no race/ethnicity group demonstrated statistically significant joinpoints, and most mortality trends were best described by single linear segments. AAMR increased from 21.44 to 28.28 per 100,000 among White individuals (APC/AAPC 6.27%, 95% CI 3.27–9.15, p<0.001), from 16.52 to 22.50 among Black individuals (APC/AAPC 7.19%, 95% CI 0.31–14.65, p=0.042), from 7.83 to 10.82 for Asian/Pacific Islander individuals (APC/AAPC 7.87%, 95% CI 2.47–13.63, p=0.004), 11.84 to 15.14 among Hispanic individuals (APC/AAPC 5.52%, 95% CI 1.10–10.00, p=0.011), and 13.74 to 16.55 among American Indian/Alaska Native individuals, though there was no statistical significance for the latter group (AAPC 5.27%, 95% CI −1.32 to 12.31, p=0.126).

### 3.3 Age-Stratified Trends

From 1999 to 2020, crude mortality rates associated with HF, AF/AFL, and sepsis increased significantly across all adult age groups, with evidence of temporal acceleration beginning around 2012 in most strata. Among individuals aged 25–34 years, the crude mortality rate increased from 0.11 per 100,000 in 1999 to 0.30 per 100,000 in 2020. Among those aged 35–44 years, rates increased from 0.30 to 0.89 per 100,000; among ages 45–54 years, from 1.08 to 3.87 per 100,000; among ages 55–64 years, from 5.00 to 14.23 per 100,000; among ages 65–74 years, from 21.73 to 46.82 per 100,000; among ages 75–84 years, from 88.50 to 171.17 per 100,000; and among ages ≥85 years, from 368.78 to 717.77 per 100,000.

Joinpoint regression identified 2012 as a statistically significant inflection point for individuals aged 35–44, 45–54, 55–64, 65–74, 75–84, and ≥85 years, whereas individuals aged 25–34 years exhibited a single continuous increasing trend without an identifiable joinpoint (APC, 4.70%; 95% CI, 3.39%–6.14%; p<0.001; AAPC, 4.70%; 95% CI, 3.39%–6.14%).

In the pre-2012 segments, mortality increased at modest but statistically significant rates across most age groups. Among individuals aged 35–44 years, mortality rose at an APC of 3.11% per year from 1999 to 2012 (95% CI 1.08%–4.09%; p=0.022), while among those aged 45–54 years, the APC was 2.62% (95% CI 0.12%–3.84%; p=0.046). For individuals aged 55–64 years, the pre-2012 APC was 1.37% (95% CI 0.58%–2.00%; p=0.004). Among older adults aged 75–84 years and ≥85 years, pre-2012 APCs were 1.24% (95% CI 0.79%–1.60%; p<0.001) and 1.47% (95% CI 1.01%–1.84%; p<0.001), respectively. The APC for individuals aged 65–74 years during 1999–2012 was not statistically significant (APC 0.48%, 95% CI −0.06%–0.93%; p=0.071), indicating stable mortality during this earlier period.

Following 2012, mortality rates accelerated markedly across all age groups with identified joinpoints. The largest post-2012 increases were observed among middle-aged adults, with individuals aged 55–64 years experiencing an APC of 10.30% per year from 2012 to 2020 (95% CI 8.92%– 12.20%; p<0.001) and those aged 45–54 years experiencing an APC of 9.98% (95% CI 7.26%–17.22%; p<0.001). Among individuals aged 35–44 years, the post-2012 APC was 8.28% (95% CI 6.21%–13.75%; p<0.001). Older adults also demonstrated significant acceleration, with APCs of 8.48% for ages 65–74 years (95% CI 7.48%–9.79%; p<0.001), 5.83% for ages 75–84 years (95% CI 5.05%–6.88%; p<0.001), and 5.82% for ages ≥85 years (95% CI 5.05%–6.85%; p<0.001). Overall, the AAPC for the entire 1999–2020 period ranged from 2.97% (95% CI 2.75%–3.18%) among individuals aged 75–84 years to 5.36% (95% CI 4.52%–6.13%) among those aged 45–54 years, reflecting sustained and statistically significant increases in mortality across all age strata. Despite younger and middle-aged adults experiencing steeper relative annual increases, older adults bore substantially greater absolute mortality throughout the study period.

During the more recent 2018–2023 period, joinpoint analysis identified statistically insignificant joinpoints across any age group. The crude mortality rate increased from 0.19 per 100,000 in 2018 to 0.26 per 100,000 in 2023 among individuals aged 25–34 years, from 0.76 to 1.11 per 100,000 among those aged 35–44 years, from 2.76 to 4.24 per 100,000 among ages 45–54 years, from 10.72 to 14.78 per 100,000 among ages 55–64 years, from 37.15 to 49.54 per 100,000 among ages 65–74 years, from 144.41 to 183.72 per 100,000 among ages 75–84 years, and from 644.74 to 843.30 per 100,000 among ages ≥85 years. Mortality rates continued to increase significantly among individuals aged 35–44 years (APC, 10.07%; 95% CI, 3.08%–17.43%; p=0.004; AAPC, 10.07%; 95% CI, 3.08%–17.43%), 75–84 years (APC, 5.60%; 95% CI, 1.73%–9.59%; p=0.004; AAPC, 5.60%; 95% CI, 1.73%–9.59%), and ≥85 years (APC, 5.86%; 95% CI, 3.63%–8.04%; p<0.001; AAPC, 5.86%; 95% CI, 3.63%–8.04%).

Trends among individuals aged 25–34, 45–54, 55–64, and 65–74 years did not reach statistical significance although some point estimates suggested continued increase in some groups, with trends approaching significance among 45–54 (p=0.064) and 65-74 years (p=0.054). The APC was equivalent to the AAPC for all age groups during 2018–2023, reflecting the absence of inflection points over this shorter period. This period encompassed the COVID-19 pandemic, during which mortality rates associated with HF, AF/AFL, and sepsis remained elevated across age groups compared with pre-pandemic levels.

### 3.4 Sex-Stratified Trends

From 1999 to 2020, AAMR associated with HF, AF/AFL, and sepsis increased significantly among both males and females, with distinct temporal inflection points identified by joinpoint regression (**Table 4**). Among males, the AAMR increased from 12.87 per 100,000 in 1999 to 28.69 per 100,000 in 2020. Among females, the AAMR increased from 11.03 per 100,000 in 1999 to 20.19 per 100,000 in 2020. Among females, mortality increased modestly from 1999 to 2012 (APC, 0.97%; 95% CI, 0.50–1.35; P<0.001), followed by an accelerated increase from 2012 to 2020 (APC, 5.54%; 95% CI, 4.75–6.61; P<0.001), corresponding to an overall AAPC of 2.69% (95% CI, 2.46–2.91). Among males, mortality increased from 1999 to 2011 (APC, 1.39%; 95% CI, 0.91–1.81; P<0.001), with a subsequent accelerated increase from 2011 to 2020 (APC, 6.67%; 95% CI, 6.03–7.53; P<0.001), yielding a higher overall AAPC of 3.62% (95% CI, 3.42–3.84). Consistent with their higher AAMR, males also experienced steeper relative increases in mortality trends compared to females, with the joinpoint occurring one year earlier (2011 vs 2012).

**Table 4.**
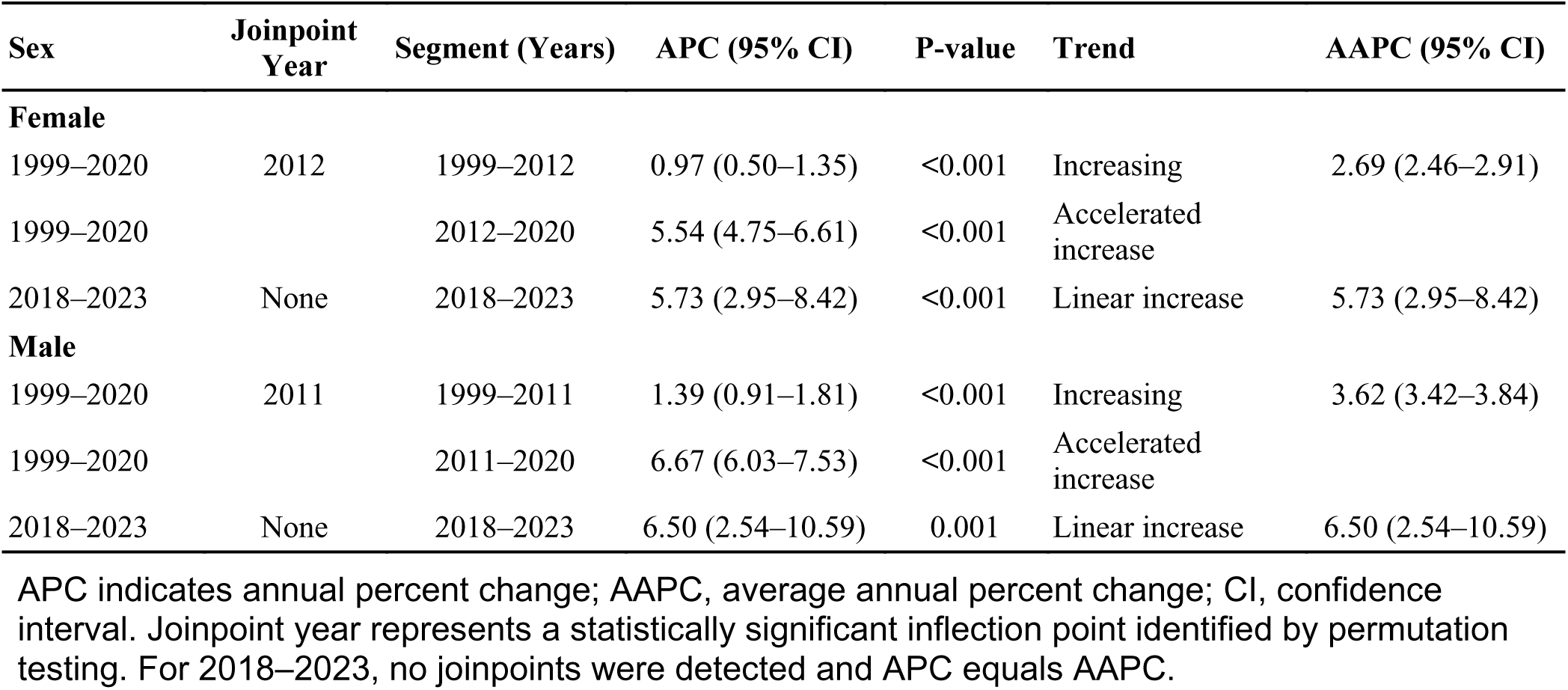
Sex-Stratified Joinpoint Trends in Age-Adjusted Mortality Rates Associated With Heart Failure, Atrial Fibrillation/Flutter, and Sepsis in the United States.

During the more recent period from 2018 to 2023, statistically insignificant joinpoints were detected for either sex. The AAMR increased from 24.03 per 100,000 in 2018 to 31.87 per 100,000 in 2023 among males and from 17.67 per 100,000 in 2018 to 22.73 per 100,000 in 2023 among females. Mortality continued to increase linearly among both females (APC/AAPC, 5.73%; 95% CI, 2.95–8.42; P<0.001) and males (APC/AAPC, 6.50%; 95% CI, 2.54–10.59; P=0.001), with males demonstrating a numerically greater magnitude of increase (Table 4).

### 3.5 Census Region Trends

From 1999 to 2020, AAMR associated with HF, AF/AFL, and sepsis increased across all United States census regions, with evidence of temporal acceleration occurring around 2010-2012 depending on region (**Table 5**). The AAMR increased from 12.00 per 100,000 in 1999 to 20.80 per 100,000 in 2020 in the Northeast, from 12.21 to 26.29 per 100,000 in the Midwest, from 11.39 to 23.41 per 100,000 in the South, and from 11.71 to 24.62 per 100,000 in the West.

**Table 5.**
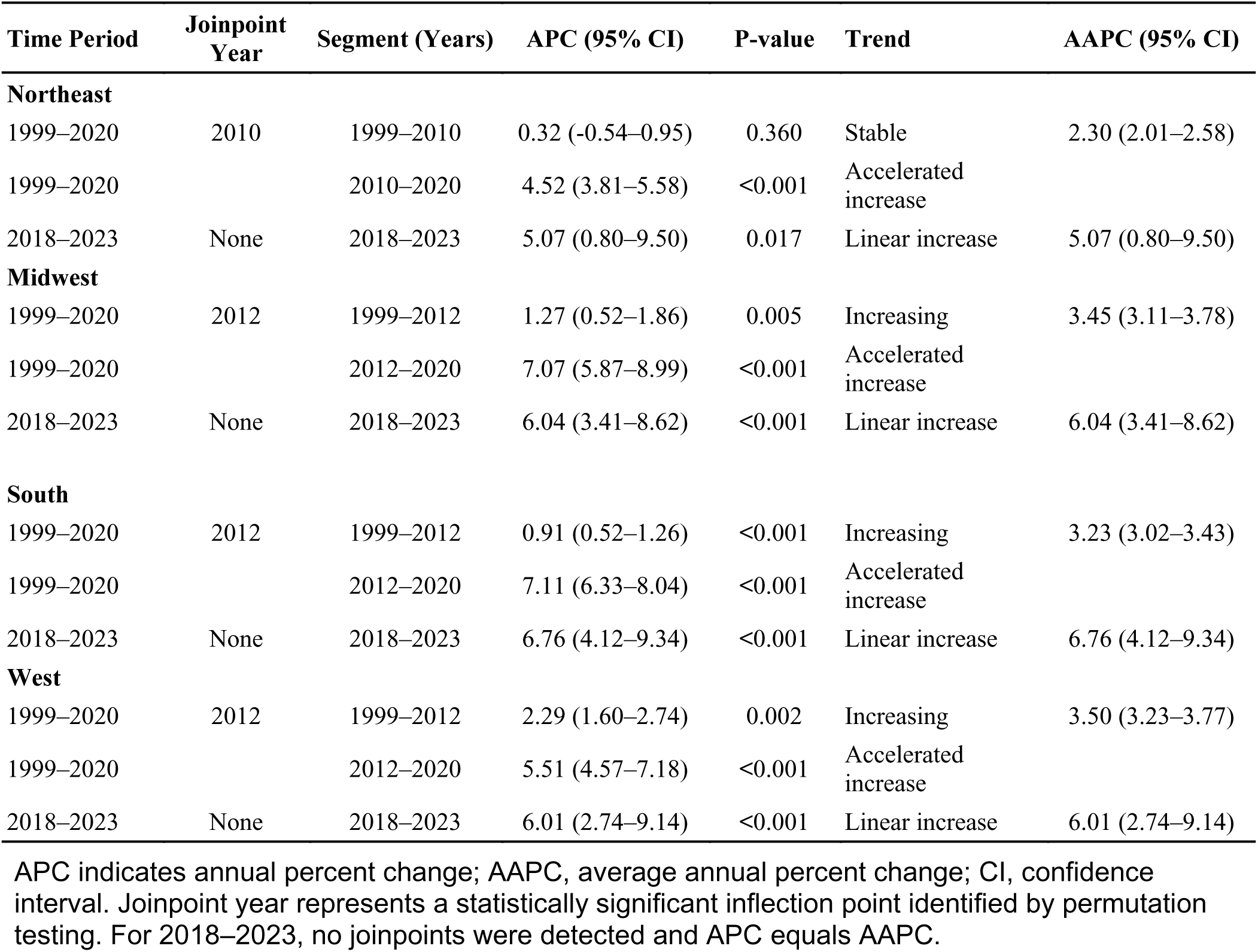
Census Region–Stratified Joinpoint Trends in Age-Adjusted Mortality Rates Associated with Heart Failure, Atrial Fibrillation/Flutter, and Sepsis in the United States.

In the Northeast, mortality rates showed no statistically significant change from 1999 to 2010 (APC 0.32%, 95% CI −0.54% to 0.95%; p=0.360), followed by a statistically significant accelerated increase from 2010 to 2020 (APC 4.52%, 95% CI 3.81%–5.58%; p<0.001; AAPC 2.30%, 95% CI 2.01%–2.58%).

In the Midwest, joinpoint regression identified 2012 as a significant inflection point, with mortality rates increasing from 1999 to 2012 (APC 1.27%, 95% CI 0.52%–1.86%; p=0.005) and accelerating thereafter from 2012 to 2020 (APC 7.07%, 95% CI 5.87%–8.99%; p<0.001; AAPC 3.45%, 95% CI 3.11%–3.78%).

Similarly, in the South, mortality rates increased from 1999 to 2012 (APC 0.91%, 95% CI 0.52%–1.26%; p<0.001), followed by a pronounced acceleration from 2012 to 2020 (APC 7.11%, 95% CI 6.33%–8.04%; p<0.001; AAPC 3.23%, 95% CI 3.02%–3.43%). In the West, a significant inflection point was also observed in 2012, with increasing rates from 1999 to 2012 (APC 2.29%, 95% CI 1.60%–2.74%; p=0.002) and an accelerated increase from 2012 to 2020 (APC 5.51%, 95% CI 4.57%–7.18%; p<0.001; AAPC 3.50%, 95% CI 3.23%–3.77%). Despite the Northeast experiencing an earlier joinpoint (2010 vs 2012) and the lowest overall AAPC (2.30%), the Midwest and South demonstrated the steepest post-joinpoint accelerations (7.07% and 7.11%, respectively), reflecting persistent regional disparities in cardiovascular health outcomes. The Midwest maintained the highest absolute mortality burden throughout the study period despite having a mid-range AAPC (3.45%), while the West exhibited the highest overall AAPC (3.50%) despite more modest post-2012 acceleration (5.51%).

During the more recent 2018–2023 period, joinpoint analysis identified no statistically significant inflection points across any census region. The AAMR increased from 17.68 per 100,000 in 2018 to 22.13 per 100,000 in 2023 in the Northeast, from 22.02 to 29.10 per 100,000 in the Midwest, from 19.97 to 26.82 per 100,000 in the South, and from 21.68 to 27.79 per 100,000 in the West. Mortality rates increased linearly in the Northeast (APC/AAPC 5.07%, 95% CI 0.80%–9.50%; p=0.017), Midwest (6.04%, 95% CI 3.41%–8.62%; p<0.001), South (6.76%, 95% CI 4.12%–9.34%; p<0.001), and West (6.01%, 95% CI 2.74%–9.14%; p<0.001). For all regions during 2018-2023, the APC was equivalent to the AAPC, reflecting the absence of joinpoints over this shorter interval, which encompassed the COVID-19 pandemic. The South demonstrated the highest 2018–2023 APC (6.76%), while the Northeast showed the most modest increase (5.07%), maintaining the regional hierarchy observed during the 1999–2020 period (**Table 5**).

## 4. Discussion

This study characterizes national trends in mortality involving HF, AF/AFL, and sepsis across demographic groups and United States regions from 1999 to 2023. AAMR more than doubled from 11.79 to 23.87 per 100,000 between 1999 and 2020, with a critical inflection point around 2012 (**Table 1**). Following this inflection, mortality accelerated from 1.29% to 6.42% annually during 2012–2020, yielding an overall AAPC of 3.21% for the full 1999–2020 period (**Table 1**). This acceleration persisted through 2023 at 6.11% per year without stabilization. To our knowledge, this is the first study describing long-term population-level mortality trends associated with concurrent HF, AF/AFL, and sepsis.

The concurrent triad demonstrates distinct and worse mortality trends compared to pairwise combinations. HF-AF/AFL mortality increased from 8.2 to 24.3 per 100,000 population (1999–2024) with acceleration from 2011–2018, then stabilized in 2021-2024 [8]. Similarly, AF-related HF deaths showed AAMR increasing from 8.15 to 20.43 per 100,000 population (1999–2020) with acceleration from 2011 onwards [9]. Sepsis-cardiovascular mortality initially declined from 38.9 to 35.4 per 100,000 (1999–2019), then reversed with increases from 2012 onwards [10]. Another study found sepsis-cardiovascular AAMR declining from 65.7 to 58.8 per 100,000 (1999–2013), then increasing to 74.3 per 100,000 in 2022 (APC 3.23%, 95% CI: 2.18%–5.40%) [11]. In contrast to these dyad patterns where cardiovascular-sepsis mortality initially declined before reversing, and HF-AF/AFL mortality ultimately stabilized during 2021–2024, the concurrent triad demonstrates sustained 6.11% annual increases through 2023 without any period of decline or stabilization, suggesting patients with all three conditions represent a uniquely vulnerable population that has not benefited from recent improvements in cardiovascular or sepsis care.

Each component of the triad independently increases the risk of developing others, creating a mutually reinforcing pathophysiology that explains the sustained mortality acceleration observed in this population. Sepsis survivors have a 1.65-fold increased risk of developing congestive heart failure that persists up to 5 years post-hospital discharge, mediated through acute cardiac dysfunction from inflammation, mitochondrial dysfunction, and chronic effects from gut barrier dysfunction allowing bacterial translocation [12, 13, 14]. Conversely, HF predisposes patients to sepsis, with 23.6% of HF patients hospitalized for infections developing sepsis, mediated by venous congestion leading to bowel edema, lymphatic dysregulation, and impaired immunity [15, 16]. Furthermore, sepsis triggers AF in 8-23% of patients through inflammatory cytokines and ion channel remodeling [17, 18, 19, 20]. Meanwhile, AF/AFL predisposes patients to HF through tachycardia-induced cardiomyopathy, while HF promotes atrial arrhythmias through elevated left atrial pressure causing fibrosis and slowed conduction [21, 22, 23, 24]. Therefore, each condition simultaneously worsens outcomes while creating the substrate for others, establishing a self-perpetuating cycle that could explain the sustained mortality acceleration.

The 2012 inflection point observed in overall mortality trends temporally coincides with the Hospital Readmissions Reduction Program (HRRP), implemented with financial penalties beginning in 2012. Multiple analyses demonstrate temporal association between HRRP and increased 30-day and 1-year post-discharge mortality, though causality remains debated [25, 26]. Hospitals adopted quality improvement initiatives focused on discharge planning and transitional care [27]. While these standard 7–14-day follow-up appointments and post-discharge phone calls may be beneficial for general HF patients, it may be fundamentally insufficient for patients presenting with the concurrent triad, who require simultaneous specialized monitoring for arrhythmia burden, volume status, anticoagulation management and infection recurrence [28, 29]. However, optimal post-discharge management for this high-risk population remains undefined, warranting further investigation.

Middle-aged adults (45-64 years) experienced steepest post-2012 acceleration (APC 9.98-10.30%), nearly double those aged >85 years (5.82%). This pattern may reflect rising cardiometabolic burden and attenuated risk factor control beginning 2010–2012 [30, 31]. Among diabetic adults, obesity increased from 46.9% in 1999-2002 to 58.1% in 2015–2020, while glycemic control initially improved to 51.8% in 2007-2010, then declined to 48% in 2015–2020. The proportion of adults with diabetes achieving control of all three risk factors (blood pressure, glycemic and lipids) peaked at 21% in 2011–2014 before declining to 18.5% in 2015–2020, temporally coinciding with the observed mortality inflection [30]. Traditional modifiable risk factors conferred greater relative risk in younger adults (<55 years) compared to older adults (>75 years), explaining 75% of HF risk in younger adults compared to 53% in older adults [31].

Our study also reported significant sex-based disparities in mortality trends (Table 4). Compared to females, males demonstrated higher AAMR throughout the study period, with an earlier joinpoint (2011 vs 2012) and higher AAPC (3.62% vs 2.69%), reflecting greater baseline burden across all three triad components. Males had higher HF mortality rates, higher age-standardized AF/AFL mortality rates, higher sepsis hospitalization rates, 56% higher age-adjusted AF incidence, and higher mortality rates secondary to septic shock [32, 33, 34]. Additionally, males have higher age-standardized cardiovascular disease mortality rates relating to ischemic heart disease, because of elevated low-density lipoprotein (LDL) cholesterol and higher tobacco use [35]. Furthermore, the earlier joinpoint noted in this group is likely due to greater baseline vulnerability to systemic healthcare changes occurring around the same time. Females, on the other hand, have more cardioprotective effects at younger ages due to the presence of estrogen [36].

In terms of racial/ethnic disparities, although Black populations had slightly higher mortality rate compared to White populations in 1999, the latter group experienced steeper post-2012 acceleration and surpassed Black populations by 2020. This trend in White populations may reflect the impact of the opioid epidemic which is associated with increased cardiovascular risk, obesogenic environment and higher baseline incident AF risk [37, 38, 39]. In contrast, Black populations have lower AF risk, but disparities persist due to social determinants preventing them from benefiting from improvements. In fact, this disparity dissipated when social determinants were controlled for [40]. Trends in Hispanic populations are attributable to acculturations, healthcare access barriers and poor risk factor control [41, 42, 43]. Asian/Pacific Islander populations had lowest overall AAMR but demonstrated the highest 2018–2023 AAPC (7.87%), likely reflecting heterogeneity masked by aggregation. Native Hawaiian/Pacific Islander adults experience 1.5-fold higher cardiovascular mortality than other Asian adults [44]. Additionally, recent data show ischemic heart disease mortality in Asian Indian adults now exceeds or equals those of White individuals [45].

Regional disparities revealed steepest post-2012 acceleration for the Midwest and South, compared to the Northeast, like prior studies demonstrating geographic clustering of poor cardiovascular health, rising cardiometabolic burden and socioeconomic disadvantage in these regions [46, 47, 48]. The Northeast had an earlier inflection point in 2010, yet during 2018-2023, all four regions demonstrated continued significant increases without plateau. Notably, the sustained 6.11% annual increase through 2023 suggests that COVID-19 accelerated pre-existing trends rather than creating new ones. COVID-19 likely triggered viral sepsis, new-onset AF and HF exacerbations, all while disrupting healthcare, which further accelerated mortality [49, 50, 51, 52].

## 5. Clinical Implications

The findings of this study have important clinical implications. First, a multidisciplinary team consisting of cardiologists, nurse practitioners, community healthcare workers, and pharmacists is needed to ensure high-risk patients have comprehensive support after discharge. Post-discharge care should focus on medication titration, psychosocial support, comorbidity management, and addressing precipitating factors. The standard 7–14-day follow-up visit may be insufficient for these patients with concurrent triads therefore serial assessments should be done to ensure clinical stability. Second, risk-stratification should extend beyond elderly populations, and should also focus on middle-aged adults, males, high-risk racial/ethnic populations, and geographic groups to ensure targeted prevention intervention is executed. Third, patients presenting with all three conditions require careful acute therapeutic management. Judicious fluid resuscitation is appropriate in septic HF patients; however, anticoagulation administration requires risk-benefit assessment as the risk of bleeding may be increased in septic patients [53,54]. AF/AFL management should be tailored to symptom burden and hemodynamic stability [55].

## 6. Limitations

One main limitation of this study is that the multiple-cause-of-death approach is unable to determine the causal pathway among HF, AF/AFL, and sepsis, making it challenging to identify the precipitating factors for clinical intervention. Additionally, changing sepsis definitions may result in coding inconsistencies. Besides that, there is currently no validation studies for the concurrent presence of all three conditions, therefore the accuracy of coding remains unknown. Furthermore, given that this is a population-level study, we are unable to determine individual-level causality.

## 7. Conclusion

In conclusion, mortality involving the concurrent presence of HF, AF/AFL, and sepsis has increased over the past two decades, with evidence of acceleration and no post-pandemic stabilization. These findings identify a distinct, high-risk population that has not benefited from advances in cardiovascular or sepsis care and underscore the need for integrated, equity-focused management strategies.

## Consent for publication

All authors have read the manuscript and consented for publication.

## Data Availability

The CDC Wonder database was utilized to extract age-adjusted mortality rates (AAMR) per 100,000 for deaths listing HF, AF/AFL, and sepsis. Although all extracted information is presented in the manuscript, any further information could be obtained by written to the corresponding author.

